# Comparative analysis of immune-associated genes in COVID-19, cardiomyopathy and venous thromboembolism

**DOI:** 10.1101/2020.08.28.20184234

**Authors:** Grant Castaneda, Abby C Lee, Wei Tse Li, Chengyu Chen, Jaideep Chakladar, Eric Y. Chang, Weg M. Ongkeko

## Abstract

As of 28 August 2020, there have been 5.88 million Coronavirus Disease 2019 (COVID19) cases and 181,000 COVID-19 related deaths in the United States alone. Given the lack of an effective pharmaceutical treatment for COVID-19, the high contagiousness of the disease and its varied clinical outcomes, identifying patients at risk of progressing to severe disease is crucial for the allocation of valuable healthcare resources during this pandemic. Current research has shown that there is a higher prevalence of cardiovascular comorbidities amongst patients with severe COVID-19 or COVID-19-related deaths, but the link between cardiovascular disease and poorer prognosis is poorly understood. We believe that pre-existing immune dysregulation that accompanies cardiovascular disease predisposes patients to a harmful inflammatory immune response, leading to their higher risk of severe disease. Thus, in this project, we aim to characterize immune dysregulation in patients with cardiomyopathy, venous thromboembolism and COVID-19 patients by looking at immune-associated gene dysregulation, immune infiltration and dysregulated immunological pathways and gene signatures.

## 1. Introduction

In December 2019, widespread infection by severe acute respiratory syndrome coronavirus 2 (SARS-CoV-2), was reported in Wuhan, Hubei Province, China [1]. Since then, SARS-CoV-2 has spread rapidly throughout China and across the world and was declared a global pandemic by the World Health Organization (WHO) on March 11, 2020 [2]. Infection by SARS-CoV-2 causes Coronavirus Disease 2019 (COVID-19). Clinical outcomes vary widely amongst patients [3-5]. Common symptoms of COVID-19 include fever, cough, fatigue, slight dyspnoea, sore throat, headache, conjunctivitis and gastrointestinal issues [6,7]. Patients with more severe cases of COVID-19 experience respiratory failure and pneumonia that may be deadly [8]. Current research suggests that patients with existing comorbidities, such as hypertension, cardiovascular disease, diabetes, or obesity are more likely to develop severe COVID-19 [9-11]. COVID-19 has also been known to induce myocardial injury, arrhythmia, acute coronary syndrome and venous thromboembolism (VTE) [12-14]. Development of cardiovascular damage has been attributed to cytokine storms triggered by SARS-CoV-2 infection that can cause multi-organ damage [15,16]. In addition, COVID-19 patients experience coagulation abnormalities, which may lead to increased risk of thromboembolic events [17]. In all, current research suggests a link between cardiovascular disease and COVID-19. However, how cardiovascular disease results in poorer COVID-19 prognosis and higher mortality rates remains unclear.

Cardiomyopathy is a cardiovascular disease where the heart muscles become inflamed, making it difficult for the heart to pump blood throughout the body. One of the most common forms of cardiomyopathy is dilated cardiomyopathy (DCM), where the ventricles of the heart are enlarged [18]. The primary cause of DCM is unclear. However, DCM can be caused by viral infections that target the heart muscles and can thereby cause heart attacks or coronary artery disease. When DCM is induced by a heart attack or coronary artery disease it is known as ischemic cardiomyopathy (ICM). By promoting an inflammatory milieu and fibrosis, cytokines and inflammasomes play a role in cardiomyopathy pathogenesis, which suggests a potential connection between cardiomyopathy and COVID-19 [19-21].

VTE includes both deep vein thrombosis (DVT), where a blood clot forms in a deep vein, and pulmonary embolism (PE), where a clot dislodges and travels to the lungs via the bloodstream, blocking pulmonary circulation. It is well-established that the immune system plays a role in DVT pathogenesis, and restriction of venous blood flow leads to the recruitment of neutrophils, monocytes and platelets [22-24]. Given that higher levels of monocytes and neutrophils have been observed in COVID-19 patients requiring ICU hospitalization, it is possible that such pre-existing immune dysregulation in COVID-19 VTE patients makes them more likely to progress to severe disease [25-27].

In this project, we aim to characterize dysregulation of the immune landscape in patients with cardiomyopathy, with VTE and with COVID-19. We analyzed the expression of cytokine genes and inflammasome-related genes using RNA-sequencing data obtained from COVID-19 and cardiomyopathy patients We also analyzed the extent of immune infiltration and the enrichment of immunological pathways and signatures. By comparing these features of the immune system, we hope to gain a more comprehensive understanding of the effects of cardiovascular disease on the immune system that leave patients more vulnerable to severe COVID-19.

## 2. Results

### 2.1 Similarities in immune-associated gene dysregulation landscape in COVID-19 and cardiomyopathy samples

We obtained gene expression sequencing data from samples of COVID-19 patients and cardiomyopathy patients and performed differential expression analysis to identify genes dysregulated between patient cohorts (edgeR, p<0.05). Gene dysregulation was determined by comparing COVID-19 samples to normal samples (from patients without COVID-19) and cardiomyopathy samples to normal samples (from patients with no major cardiovascular diseases). Cardiomyopathy samples were separated based on whether patients had ischemic or dilated cardiomyopathy. The two groups were individually compared against samples from patients with no major cardiovascular disease. Cardiomyopathy and corresponding normal samples were obtained from the left ventricle of deceased patients, while COVID-19 and corresponding normal samples were platelet samples. However, we expect to capture genetic material from circulating immune cells in both of these samples.

Only immune associated (IA) differentially expressed genes were retained for further analysis, in order to compare the immune profile of patients with COVID-19 to that of patients with cardiomyopathy. We found that there was significant overlap between COVID-19 patient IA gene expression and that of both ischemic and dilated cardiomyopathy patients. About half of the IA genes dysregulated in COVID-19 are also dysregulated in either or both types of cardiomyopathy (Figure 1A). We grouped these genes into cytokine-related and inflammasome-related genes. Cytokine-related genes that are dysregulated in both cardiomyopathy patients and COVID-19 patients include chemokines (CCL3, CCL4, CXCL4, etc), interleukins or interleukin receptors (IL15, IL20RA, etc.), or genes in the TGFB family. Inflammasome-related genes include genes in the caspase family (CASP2, CASP9, etc), MAPK-related genes, and NFkB regulators (IKBKG, NFKBIA, etc.). IA gene dysregulation is very similar between dilated and ischemic cardiomyopathies. We observed that a significant number of cytokines, interleukins, TGFB-related genes, and TNF-related genes were dysregulated in either types of cardiomyopathies but not in COVID-19 (Figure 1A).

**Figure 1:**
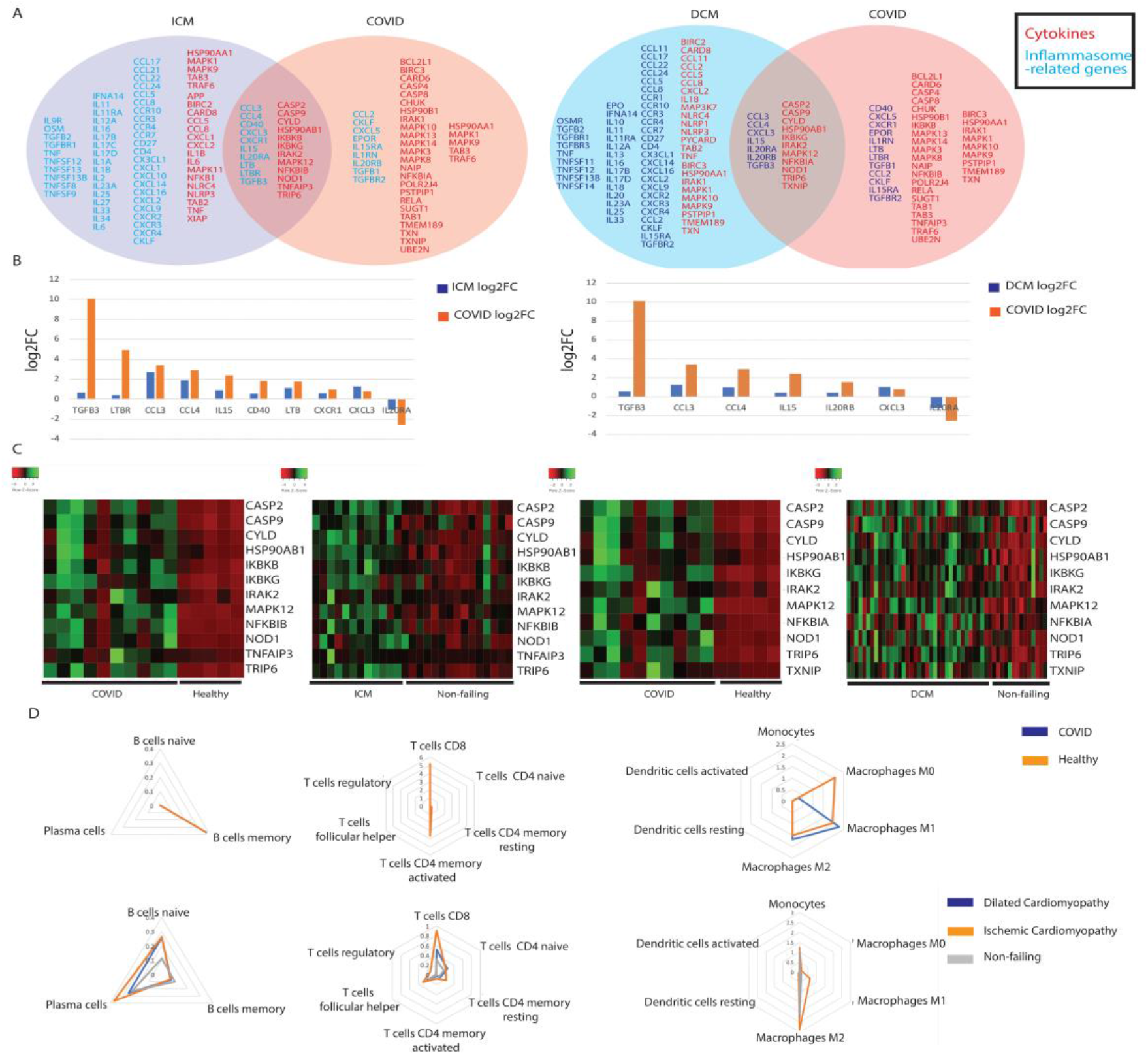
Comparing ICM, DCM and COVID-19 patients. (**A**) Summary of commonly dysregulated cytokine and inflammasome-related genes in COVID-19 and ICM/DCM patients. Cytokines are in blue and inflammasome-related genes are in red. (**B**) Barplots of the log2 fold change of significantly dysregulated cytokine genes in COVID-19 and ICM/DCM patients. (**C**) Heatmaps of inflammasome-related genes in COVID-19 and ICM/DCM patients. (**D**) Radar plots comparing immune cell infiltration between COVID-19 and ICM/DCM patients.

Interestingly, we found that most genes dysregulated in both COVID-19 and cardiomyopathy, were dysregulated to a greater degree in COVID-19 samples than in cardiomyopathy samples. This trend was observed for TGFB3, CCL4, IL15, and IL20RA in both ischemic cardiomyopathy samples vs COVID-19 samples and dilated cardiomyopathy samples vs. COVID-19 samples (Figure 1B). Furthermore, the expression of these dysregulated genes appeared to be very similarly dysregulated in COVID-19 samples and in corresponding healthy samples, with similar levels of gene expression for individual patients in each group and a distinct upregulation trend in COVID-19 samples (Figure 1C). In contrast, these genes’ expression in cardiomyopathy samples and corresponding healthy samples are only sometimes similar, without overwhelming differences in expression levels between the two cohorts (Figure 1C). Therefore, we believe that these dysregulated genes are dysregulated to a greater degree in COVID-19 than in cardiomyopathy.

The inflammasome-associated genes dysregulated in both COVID-19 and cardiomyopathy are upregulated in both conditions (Figure 1C), suggesting that COVID-19 and cardiomyopathy may upregulate inflammation through similar pathways.

### 2.2 Comparison of immune cell population abundance in COVID-19 vs. cardiomyopathy patients

Using Cibersortx, we inferred the percent composition of 22 different immune cell types using the gene expression profile of bulk RNA-sequencing data. We discovered that the levels of T cells and B cells were unchanged in healthy vs. COVID-19 patients (Figure 1D). The most noticeable change in immune cell abundance occurs in macrophages for COVID-19 patients, where M0 macrophages levels were dramatically reduced, and M1 and M2 macrophages levels were slightly increased (Figure 1D). Both ischemic and dilated cardiomyopathy elicited greater immune cell abundance changes than COVID-19 did, with the changes being more pronounced for ischemic myocardiopathy. The levels of M1 and M2 macrophages also increased in ischemic myocardiopathy, similar to what was observed for COVID-19 (Figure 1D). The levels of T and B cell subtypes changed more dramatically in ischemic and dilated myocardiopathy than in COVID-19. In summary, the levels of inflammatory macrophages increased for both cardiomyopathy and COVID-19 patients, while the levels of other immune cell types did not correlate between the two conditions.

### 2.3 Evaluation of canonical pathways correlated with genes dysregulated in both COVID-19 and cardiomyopathy

To obtain a deeper understanding of the functions of the dysregulated genes in COVID-19 and cardiomyopathy, we correlated IA gene expression to the dysregulation of canonical biological pathways, obtained from the Molecular Signatures Database (MsigDB) [28]. We analyzed genes that are dysregulated in both COVID-19 and cardiomyopathy to assess if they may dysregulate the common pathways in the two conditions. IRAK2, upregulated in both COVID-19 and ischemic cardiomyopathy, was associated with the upregulation of the FCER1 and TP63 pathways, both of which are associated with inflammation and immune activation (Figure 2A) [29,30]. IRAK2 is a promoter of NFkB signaling [31]. CASP2, also upregulated in COVID-19 and cardiomyopathy, is associated with the downregulation of IFIH, which is capable of recognizing viruses and inducing inflammation [32]. Finally, CYLD is correlated with multiple identical pathways for both COVID-19 samples and ischemic cardiomyopathy samples. It is also upregulated in both COVID-19 and cardiomyopathy and was found to correlate with the activation of FGFR2, an important promoter of inflammation [33], and TXA2, a well-known gene that is upregulated in platelets (Figure 2A) [34]. CYLD itself is an inhibitor of inflammation [35]. Since the majority of correlations are between IA genes and pro-inflammatory pathways and signatures, we hypothesize that CYLD may be expressed as a response to attenuate excessive inflammation. We found that the overwhelming majority of pathways that correlated with dysregulated genes in both COVID-19 and dilated cardiomyopathy is associated with CYLD, and these pathways are primarily pro-plotting, pro-cell aggregation, and pro-inflammation (Figure 2B), which reinforces the possibility that CYLD is released in response to inflammation.

**Figure 2:**
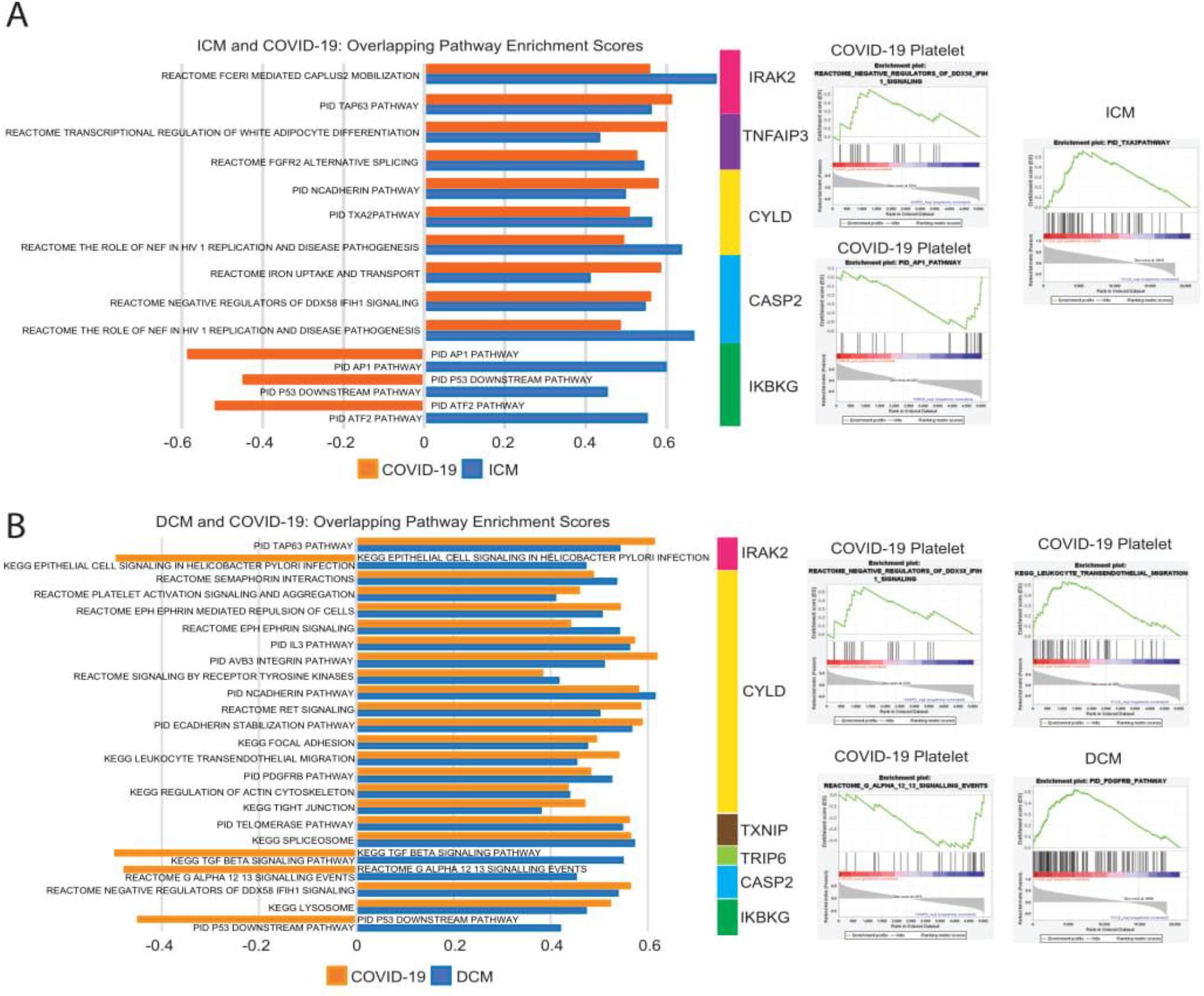
Comparing ICM, DCM and COVID-19 patients. (**A**) Barplots showing direction of correlation between commonly dysregulated genes of ICM and COVID-19 patients and canonical pathways. (**B**) Barplots showing direction of correlation between commonly dysregulated genes of DCM and COVID-19 patients and canonical pathways.

### 2.4 Similarities in immune-associated gene dysregulation landscape in COVID-19 and venous thromboembolism (VTE) samples

We compared the immune landscape between COVID-19 samples and blood samples from VTE patients to find similarities in IA gene and pathway expression. VTE patients are further classified into single occurrence VTE (single VTE) and recurrent VTE. Differential expression was performed by comparing blood samples for each type of VTE against blood samples from healthy controls. Compared to the similarities in IA gene dysregulated between COVID-19 and cardiomyopathy, the similarities between COVID-19 and VTE are much less-pronounced. We identified two cytokine-associated genes that were dysregulated in both COVID-19 and single VTE (CCL4 and CD40), and one cytokine-associated gene dysregulated in both COVID-19 and recurrent VTE (CCL4) (Figure 3A). CCL4 is a recruiter of immune cells, including macrophages, monocytes, and T-cells [36], suggesting that both COVID-19 and VTE may experience increased recruitment of inflammatory immune cells. The upregulation of CCL4 is much greater in COVID19 than in VTE, however (Figure 3B).

**Figure 3:**
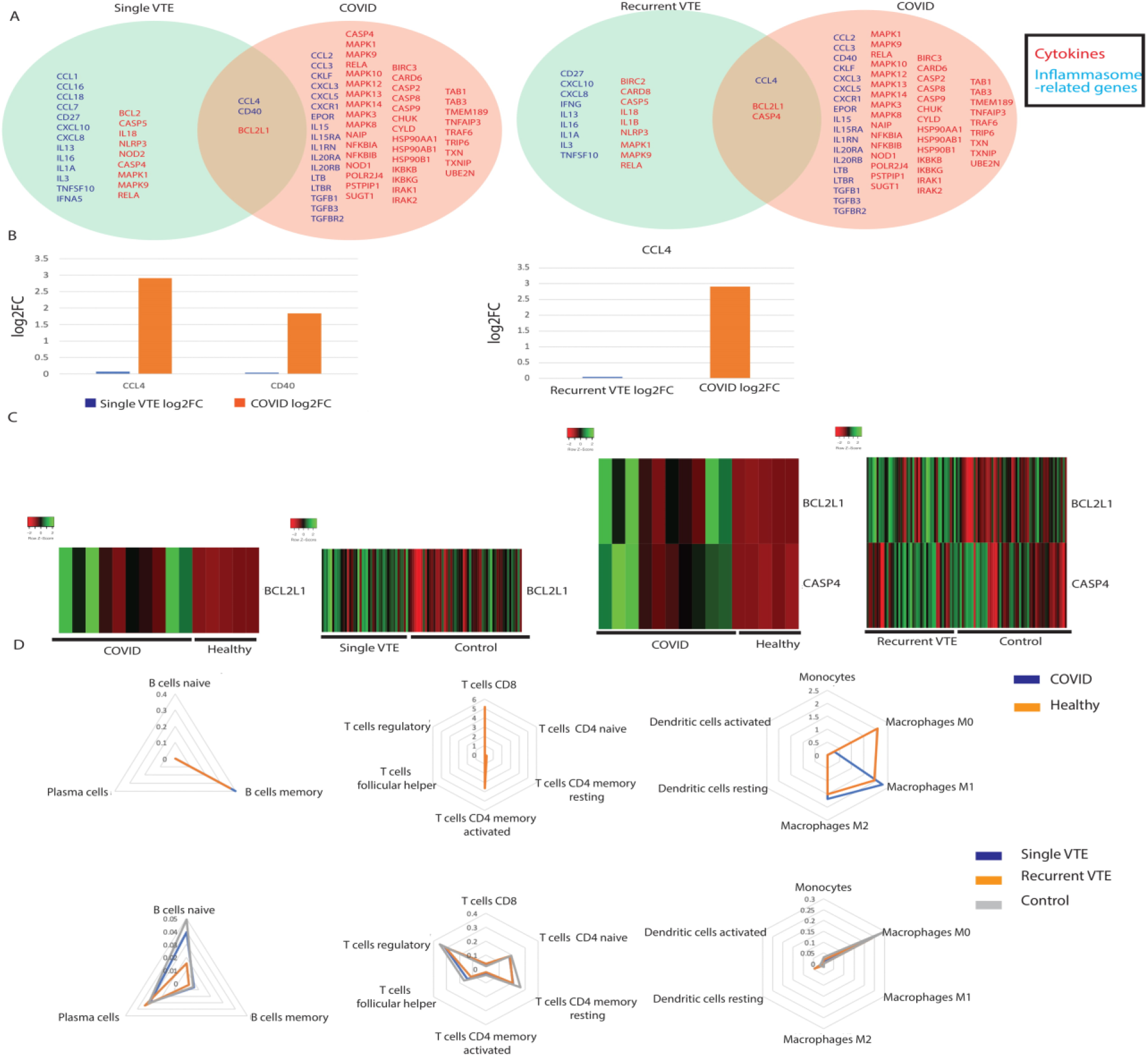
Comparing single VTE, recurrent VTE and COVID-19 patients. (**A**) Summary of commonly dysregulated cytokine and inflammasome-related genes in COVID-19 and single/recurrent VTE patients. Cytokines are in blue and inflammasome-related genes are in red. (**B**) Barplots of the log2 fold change of significantly dysregulated cytokine genes in COVID-19 and single/recurrent VTE patients. (**C**) Heatmaps of inflammasome-related genes in COVID-19 and single/recurrent VTE patients. (**D**) Radar plots comparing immune cell infiltration between COVID-19 and single/recurrent VTE patients.

Two inflammasome-related genes were found to be dysregulated in VTE and COVID-19, including BCL2L1 and CASP4. Both single and recurrent VTE share the same set of dysregulated inflammasome-related genes with COVID-19. BCL2L1 is known to be highly upregulated in inflamed tissue [37], and it is upregulated in COVID-19 and both single and recurrent VTE (Figure 3C). CASP4 directs the noncanonical upregulation of inflammasomes [38]. Interestingly, it is upregulated in both COVID-19 and recurrent VTE but downregulated in single VTE (Figure 3C), which potentially suggests that the gene could contribute to the development of recurrent VTE.

### 2.5 Comparison of immune cell population abundance in COVID-19 vs. VTE patients

Comparing immune cell population abundance between COVID-19 and VTE patients, we found that naïve B-cells were dramatically reduced in abundance in VTE patients, which is accompanied by a slight increase in activated plasma B cells (Figure 3D). This may be a sign of adaptive immune activation.

### 2.6 Evaluation of canonical pathways correlated with genes dysregulated in both COVID-19 and VTE

We adopted a similar approach as described for COVID-19 vs. cardiomyopathy to examine canonical pathways correlated with dysregulated genes in both COVID-19 and VTE. BCL2L1 and CASP4 are the only genes dysregulated in both COVID-19 and VTE and also correlated with the same pathways in both patient cohorts (Figure 4A,B). BCL2L1 is upregulated in COVID-19 and both VTE cohorts (Figure 4C). However, the direction of correlation to pathways was the complete opposite between COVID-19 and recurrent VTE (Figure 4D, E). The high strength of correlation for each cohort suggests that BCL2L1 is involved in both COVID-19 and recurrent VTE but may function in opposite ways. On the other hand, CASP4 expression was correlated with over 10 pathways in the same direction for both COVID-19 and recurrent VTE (Figure 4D). It is also upregulated in both COVID-19 and recurrent VTE (Figure 4C). The pathways correlated with CASP4 ranges from immune related (antigen processing and cross presentation) to general metabolism-related (ABC transporter, oxidative phosphorylation). Therefore, while it is likely that CASP4 functions similarly in COVID-19 and recurrent VTE. The precise role of CASP4 requires further investigation.

**Figure 4:**
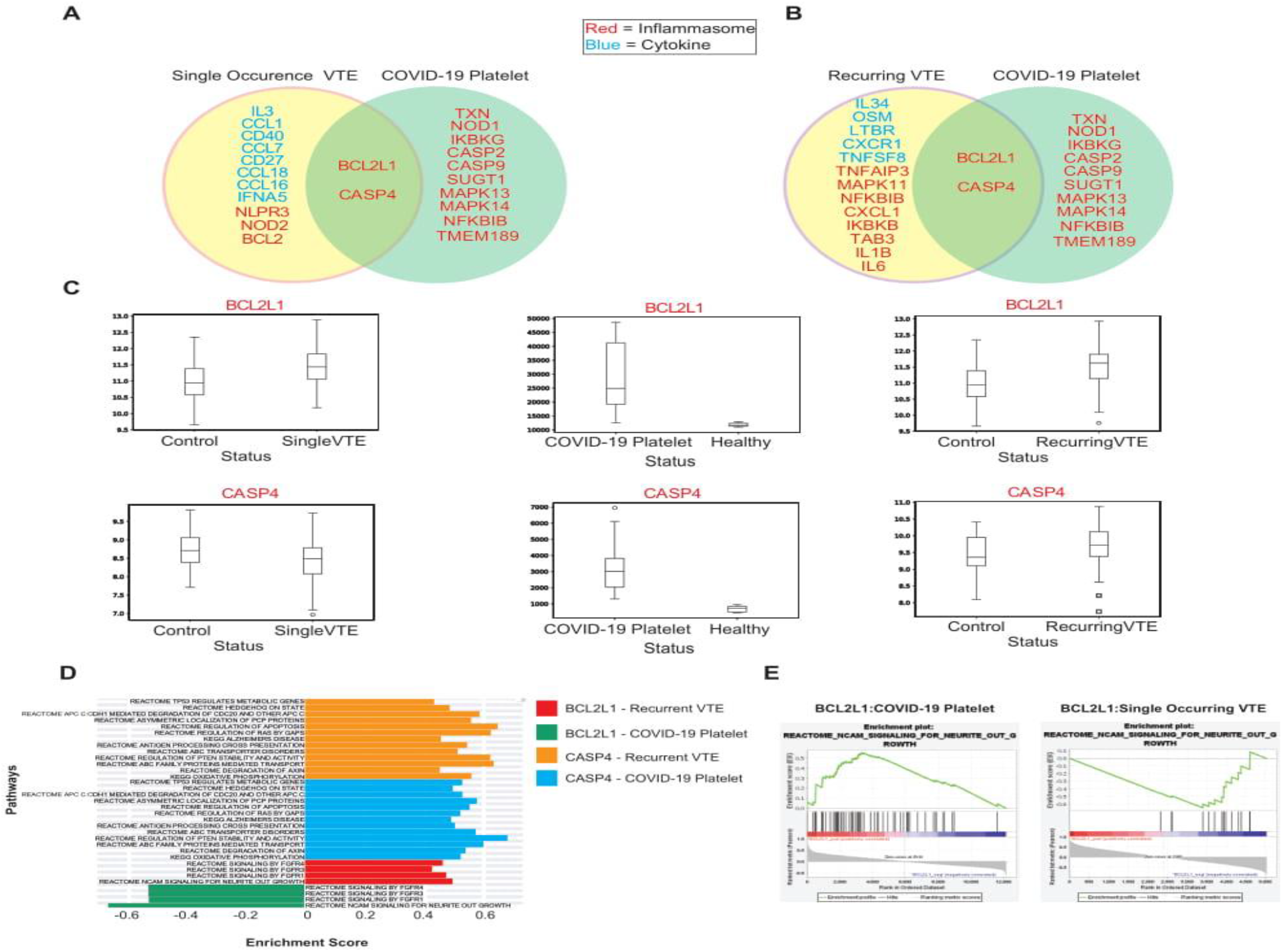
Comparing single VTE, recurrent VTE and COVID-19 patients. (**A**) Summary of common dysregulated genes correlated with pathways in single VTE and COVID-19. (**B**) Summary of common dysregulated genes correlated with pathways in recurring VTE and COVID-19. (**C**) Boxplots of CASP4 and BCL2L1 expression in COVID-19 and VTE cohorts. (**D**) Barplots showing direction of correlation between CASP4/ BCL2L1 and canonical pathways. (**E**) Enrichment plots showing BCL2L1’s correlation to pathways is opposite in COVID-19 and recurrent VTE patients.

## 3. Discussion

In this project, we characterized the immune landscape of cardiomyopathypatients, VTEpatients, and COVID-19 patients, drawing parallels between COVID-19 and immune dysregulation mediated by cardiovascular disease. The overlap of numerous dysregulated IA genes between COVID-19 patients and cardiomyopathy patients not only suggests that these patients possess similar inflammatory environments but could also explain why COVID-19 patients with cardiovascular disease have higher mortality rates. Of the four genes (TGFB3, CCL4, IL15, and IL20RA) that were more severely dysregulated in COVID-19 samples compared to cardiomyopathy patients, two have been noted to be dysregulated in COVID-19 patients: proinflammatory cytokine CCL4 was found to be highly expressed in the bronchoalveolar lavage fluid of COVID-19 patients [39], and IL-15 modulates inflammation and plays a role in viral clearance [40,41]. We also found that both cardiomyopathy and COVID-19 patients had elevated levels of inflammatory macrophages, while T and B cell levels changed more significantly in dilated and ischemic cardiomyopathy patients compared to COVID-19 patients. This points to a more robust innate immune response in COVID-19 patients, which is plausible given that research has shown that an excessively inflammatory innate immune response coupled with an impaired adaptive immune response may lead to tissue damage in COVID-19 patients [42,43]. On the contrary, the elevated levels of T and B cells in cardiomyopathy patients indicates a stronger adaptive immune response, which is now being considered an increasingly important factor in the pathogenesis of cardiovascular disease [44-46].

We analyzed overlapping gene expression pathway dysregulation between cardiomyopathy and COVID-19 patients in order to compare IA profiles between the two patient cohorts. The overlapping genes that displayed significant canonical pathway dysregulation between the cardiomyopathy and COVID-19 cohorts are CYLD, CASP2, IKBKG and IRAK2. CASP2 and IRAK2 are of particular interest due to their involvement in inflammation. CASP2 is a pro-inflammatory gene and IRAK2 promotes NFkB, which ultimately causes inflammation. Both genes are upregulated in DCM, ICM and COVID-19, suggesting that DCM and ICM patients are more susceptible to a worse inflammatory response to COVID-19. Both IRAK2 and CASP2 are directly associated with the dysregulation of inflammation pathways. For instance, IRAK2 is directly associated with the TAP63 pathway in both ICM and DCM versus COVID-19 groups, indicating that one of the effects of IRAK2 in COVID-19 and cardiomyopathy may be increasing inflammation. Similarly, CASP2 is directly associated with the IFIH1 signaling pathway, which is known to be pro-inflammation. Overall, the genes that overlap between cardiomyopathy and COVID-19 appear to act in a pro-inflammatory manner. Increased inflammatory pathway dysregulation may indicate a reason why cardiovascular disease patients experience much worse clinical outcomes, as several studies have shown a worse inflammatory response correlates with severity and death in COVID-19 [47].

Exploring the immune-association gene dysregulation landscape relationship between VTE and COVID-19 revealed overlapping differentially expression of CCL4, CD40, BCL2L1 and CASP4. Of the cytokines CCL4 and CD40, only CCL4 is commonly dysregulated in single occurring VTE, recurring VTE and COVID-19 patients. CCL4 has been shown to be upregulated in COVID-19 patients [48] and in patients who develop cardiovascular disease [49]. Therefore, the upregulation of CCL4 in both VTE and COVID-19 may suggest a mechanism by which some COVID-19 patients develop cardiovascular disease post-infection. The inflammasome-related genes dysregulated in both the VTE and COVID-19 cohorts are BCL2L1 and CASP4. BCL2L1 and CASP4 expression are closely tied to inflammatory caspases. CASP4 itself is an inflammatory caspase and it promotes the secretion of pro-inflammatory cytokines [50]. Conversely, BCL2L1 inhibits caspase release. With both of these genes being upregulated in VTE and COVID-19, future analysis must be done to examine how these genes function differently in VTE and COVID-19. BCL2L1 and CASP4, both inflammasome-related genes, stood out as the most interesting genes as both displayed significant canonical pathways in both VTE groups, single occurring and recurrent VTE, and COVID-19. However, only in recurring VTE did both BCL2L1 and CASP4 display overlapping canonical pathways with COVID-19, while single occurring VTE only had BCL2L1 with overlapping canonical pathways with COVID-19. Most notably, in both COVID-19 and single occurring VTE, BCL2L1 expression is negatively correlated to canonical pathway expression, but in recurring VTE they are positively correlated. Thus, we may deduce that a main difference between single and recurrent VTE is the utilization of BCL2L1. The downregulation of BCL2L1 may increase the risk of COVID-19. due to BCL2L1-mediated inhibition of pro-inflammatory caspases. CASP4, on the other hand, only has overlapping significant canonical pathways with COVID-19 in the recurrent VTE cohort. From these overlapping canonical pathways, we could not find a common trend like there was in BCL2L1. However, we could deduce from these pathways that CASP4 functions similarly in both recurrent VTE and COVID-19.

In conclusion, our study demonstrates that cardiomyopathy and VTE patients display a significant overlap of inflammasome gene expression profiles with that of COVID-19 patients. This may explain why patients with cardiovascular disease are more likely to develop severe COVID-19, and tend to have poorer clinical outcomes [51,52]. Furthermore, we have found that patients with cardiomyopathy display more similar immune dysregulation to COVID-19 patients than VTE patients, which may indicate that COVID-19 patients who have cardiomyopathy are at higher risk of severe COVID-19 than COVID-19 patients with VTE. Our findings also suggest that investigating the relationships between different cardiovascular diseases and COVID-19 disease severity and mortality may be meaningful, and can offer greater insight into COVID-19 immune dysregulation. However, we recognize our study has several limitations. We had limited COVID19 platelet data, specifically normal patients. Thus, our differential expression analysis may have been impacted, and the statistical power of our analysis is reduced. However, the direction of dysregulation of many of the genes we identified were consistent with existing literature. Additionally, we used platelet data, instead of blood samples. In order to validate our results, *in vitro* and *in vivo* experiments can be done in the future. Despite these limitations, we believe our study advances our understanding of the relationship between cardiovascular disease and COVID-19.

## 4. Methods and Materials

### Downloading Data

RNA-sequencing data was obtained from the following datasets: GSE116250, GSE19151 and SRP262885 (8) [53-55]. GSE116250, provided by Sweet et. al, consists of RNA-sequencing of human left ventricular samples from 14 patients with no major cardiac history (non-failing), 37 patients with dilated cardiomyopathy and 13 patients with ischemic cardiomyopathy. The mean age of patients with dilated and ischemic cardiomyopathy was 49 and 56 years respectively. GSE19151, provided by Lewis et. al, consists of high-throughput sequencing of whole blood samples from 63 healthy controls, 23 patients with single event VTE, 17 patients with recurrent event VTE on warfarin. The mean age of patients with single VTE and recurrent VTE was 43.8 and 55.7 years respectively. Lastly, SRP262885 consists of RNA-sequencing data of platelets from 10 COVID-19 subjects and 5 healthy controls. The mean age of COVID-19 patients and healthy controls is 51.1 and 43.8 respectively.

### Differential Expression

Differential expression analysis was employed to compare the expression of genes between diseased and normal individuals. For the cardiomyopathy and venous thrombosis cohorts, a Kruskal-Wallis analysis test was used to determine the differentially expressed genes, and genes with *p* < 0.05 were considered significantly dysregulated. Differential expression was applied to the COVID-19 platelet data to determine the genes that were differentially expressed (*p* < 0.05).

### GSEA

To correlate gene expression to immune-associated signatures, Gene Set Enrichment Analysis (GSEA) was utilized. Pathways were chosen from the C2:CP set of signatures from the Molecular Signatures Database [28]. Signatures that were significantly enriched were those with a nominal p-value < 0.05.

### CIBERSORTx

The CIBERSORTx algorithm was used to deconvolute RNA-sequencing data to estimate the infiltration levels of 22 immune cell types [56]. These immune cell types include: naïve B-cells, memory B-cells, plasma cells, CD8 T-cells, CD4 naïve T-cells, CD4 memory resting T-cells, CD4 memory activated T-cells, follicular helper T-cells, regulatory T-cells, gamma-delta T-cells, resting NK cells, activated NK cells, monocytes, M0-M2 macrophages, resting dendritic cells, activated dendritic cells, resting mast cells, activated mast cells, eosinophils, and neutrophils.

## Data Availability

RNA-sequencing data was obtained from the following datasets: GSE116250, GSE19151 and SRP262885.

https://trace.ncbi.nlm.nih.gov/Traces/sra/?study=SRP262885

https://www.ncbi.nlm.nih.gov/geo/query/acc.cgi?acc=GSE116250

https://www.ncbi.nlm.nih.gov/geo/query/acc.cgi?acc=GSE116250

## Author Contributions

Conceptualization, W.M.O; methodology, W.T.L and W.M.O.; software, N/A; formal analysis, A.C.L, G.C, and C.C; investigation, A.L. and G.C.; resources, W.M.O.; data curation, N/A; writing, A.C.L., G.C. and W.T.L.; visualization, A.L. and G.C.; supervision W.M.O.; project administration, W.M.O.; funding acquisition, W.M.O.

